# Detecting SARS-CoV-2 at point of care: Preliminary data comparing Loop-mediated isothermal amplification (LAMP) to PCR

**DOI:** 10.1101/2020.04.01.20047357

**Authors:** Marc F Österdahl, Karla A Lee, Mary Ni Lochlainn, Stuart Wilson, Sam Douthwaite, Rachel Horsfall, Alyce Sheedy, Simon D Goldenberg, Christopher J Stanley, Tim D Spector, Claire J Steves

## Abstract

**Background:** The need for a fast and reliable test for COVID-19 is paramount in managing the current pandemic. A cost effective and efficient diagnostic tool as near to the point of care (PoC) as possible would be a game changer in current testing. We tested reverse transcription loop mediated isothermal amplification (RT-LAMP), a method which can produce results in under 30 minutes, alongside standard methods in a real-life clinical setting.

**Methods:** This service improvement project piloted a research RT-LAMP method on nasal and pharyngeal swabs on 21 residents in a high dependency care home, with two index COVID-19 cases, and compared it to multiplex tandem reverse transcription polymerase chain reaction (RT-PCR). We calculated the sensitivity, specificity, positive and negative predictive values of a single RT-LAMP swab compared to RT-PCR, as per STARD guidelines. We also recorded vital signs of patients to correlate clinical and laboratory information.

**Findings:** The novel method accurately detected 8/10 PCR positive cases and identified a further 3 positive cases. Eight further cases were negative using both methods. Using repeated RT-PCR as a “gold standard”, the sensitivity and specificity of the novel test were 80% and 73% respectively. Positive predictive value (PPV) was 73% and negative predictive value (NPV) was 83%. We also observed hypothermia to be a significant early clinical sign in a number of COVID-19 patients in this setting.

**Interpretation:** RT-LAMP testing for SARS-CoV-2 was found to be promising, fast, easy to use and to work equivalently to RT-PCR methods. Definitive studies to evaluate this method in larger cohorts are underway. RT-LAMP has the potential to transform COVID-19 detection, bringing rapid and accurate testing to the point of care. This method could be deployed in mobile testing units in the community, care homes and hospitals to detect disease early and prevent spread.

## Introduction

Current diagnosis of for COVID-19 infection relies on centralised laboratory-based RT-PCR (Reverse Transcription Polymerase Chain Reaction) testing. Although PCR provides a relatively rapid result, it is limited by the bottle necks of transportation to the laboratory and the requirement to batch samples in a large run. Moreover, alternative technologies to RT-PCR requiring different reagents, and dry swabs would reduce the strain on laboratory and clinical supplies, allowing greater numbers of tests to be performed. It is abundantly clear that urgent research is needed to enable health services globally to plan resources and this research must both move rapidly from bench to bedside and be scalable and rapidly available. In light of this urgency, we present a preliminary evaluation of a novel, quick test for COVID-19 that can be implemented at the point of need.

Point-of-care (PoC) testing may be critical to enable rapid detection of disease when an outbreak is suspected. This is particularly important in community settings like care homes, where multiple vulnerable patients reside together, and COVID-19 can spread quickly if not identified early^[1]^. Older residents are at higher risk of mortality from COVID-19^[2]^, and care homes have reported significant outbreaks both in the UK^[3]^ and internationally^[4]^. However, they have limited access to laboratory diagnostic services. A rapid, point-of-care (PoC) test would allow early case identification, and implementation of increased infection control measures to prevent further spread to residents and staff, as recommended by Public Health England (PHE)^[5]^, The World Health Organization (WHO)^[1]^ and British Geriatric Society^[6]^.

To this end, we used a combination of magnetic bead viral genome capture and optimised RT-LAMP (Reverse Transcriptase Loop-Mediated Isothermal Amplification) for amplification and detection of the SARS-CoV-2 genome; targeting the ORF1a gene, to show proof of principle. The assay runs under isothermal conditions at 65°C allowing simpler and cheaper instrumentation to be used with rapid results (25 minutes from swab to result). It can be used without a hospital laboratory and is suitable for a mobile testing unit model.

## Research in Context

### Evidence before this study

A cost effective and efficient testing tool as near to the point of care (PoC) as possible for COVID-19 is vital for future swift detection to stem the pandemic. There is currently no “real world data” comparing the effectiveness of RT-LAMP to the current gold standard of RT-PCR but lab-based studies in China and the USA suggest these methods have similar theoretical efficacy.

### Added Value of this study

Magnetic bead capture and RT-LAMP amplification and testing for SARS-CoV-2 was found to be promising, easy to use and to work equivalently to RT-PCR methods in a “real world” cohort of patients in a care home. Hypothermia is a notable clinical sign in our cohort for patients with Covid-19 infection, and consideration should be given to its inclusion in diagnostic criteria as an alternative to fever

### Implications of all the available evidence

RT-LAMP can be used to aid rapid diagnosis for COVID-19 at the point of care in community settings, allowing RT-PCR to be reserved for use elsewhere. Hypothermia, in addition or contrast to fever, may also be a significant early clinical sign of COVID-19 disease, and further investigation is recommended for its use in clinical practice.

## Methods

### Study Design

The setting was an NHS high dependency care home (Category 1 Continuing Care), where an outbreak was suspected. On Day 0 (Monday 16^th^ March) 2 patients experienced fever and had other classical symptoms of COVID-19, arousing clinical suspicion. RT-PCR testing was performed on Day 1 and reported as positive on Day 2. To determine the extent of spread in the home, and protect patients and staff, on days 3 & 4 nasal and pharyngeal swabs were performed in all patients in the care home and analysed using multiplex tandem RT-PCR. On Day 4 RT-LAMP swab tests from the nose and throat were added to the testing protocol. Patients’ vital signs were noted in the 4 weeks prior to the outbreak to trace whether the start of the outbreak was prior to day 0.

### Test Methods

Isolation and barrier nursing were instituted for all patients. All patients were sampled on day 3 and day 4 using Pharyngeal (Day 3) and deep nasal (Day 4) specimens (swabs) collected which were immediately placed into sterile tubes containing 3ml of viral transport media (VTM) for RT-PCR or dry for the RT LAMP assay. Staff taking the swabs were also swabbed and were negative for SARS-CoV-2 using RT-LAMP. Samples were urgently couriered to the hospital and MicrosensDx laboratory.

The hospital performed multiplex tandem RT-PCR according to standard protocols. If Patients were positive on Day 3, Day 4 samples were not analysed, but have been stored for later analysis. The RT-LAMP method employed was the MicrosensDx RapiPrep® SARS-CoV-2 research use test. The method used magnetic bead capture to maximise the yield of target nucleic acid during sample preparation from the dry swab, which is followed by optimised reverse transcription loop mediated isothermal amplification (RT-LAMP) to amplify and detect the SARS-CoV-2 genome, targeting the *ORF1a* gene. The assay runs under isothermal conditions at 65°C allowing simpler and cheaper instrumentation which can yield results in 25 minutes on average. Results from this assay were compared to multiplex tandem PCR performed twice in the case of negative patients.

## Results

24 residents were present in the care home on Day 0. Two patients lacked capacity had no contactable NOK to inform of the project. In one patient their informant did not agree to repeated testing as a service improvement. 21 patients were included in the study. Study participants were aged between 52 and 89 years (median 76 years) and were predominantly female (70%). 2/21 died due to Covid-19, and 2/21 died from unrelated causes within 7 days of their positive test (Table 1).

**Table 1:** Baseline Characteristics of Cohort

|  |  |
| --- | --- |
| Total in Care Home | 24 |
| Included in Study | 21 |
| Age - Median | 76 years |
| - Interquartile Range | 61-81 years |
| Sex (Female) | 17 (70%) |
| Any Fever (>37.3C) in past 28 days | 16/21 |
| 7-day Mortality - Due to Covid | 2 (10%) |
| 7-day mortality - Other Causes | 2 (10%) |

Testing results are shown in Table 2. We defined cases as being RT-PCR positive on one of two tests at day 3 or 4, and negative if negative on both tests. Using this definition, 10/21 patients in the facility were COVID-19 positive (RT-PCR34). Of these 10 cases, 8 were identified with a single swab for RT-LAMP, giving a sensitivity of 80% (95% CI 44-98%) and positive predictive value of 73% (95%CI 39-94%) (Table 2). This represented an improved rate of detection compared to single swab RT-PCR both in our sample and previous estimates. Combining symptoms with RT-LAMP assessment did not improve sensitivity or specificity in this patient group.

**Table 2:** Participant results, vital signs and mortality at 7 days. Rows for cases (defined as at least one of two positive RT-PCR tests) are highlighted in orange.

|  |  |  |  | Vital signs in 7 days prior to any positive result |  |  |  |  |
| --- | --- | --- | --- | --- | --- | --- | --- | --- |
| ID | D1/3 PCR<br>(pharyngeal) | D4 PCR<br>(nasal) | D4 LAMP<br>(Nasal) | T>37.8C | T<36.0C | T>37.3C | SpO2<92% | Death |
| 1 | - | - | - |  |  |  |  |  |
| 2 | + | n/a | + | Yes |  |  | Yes |  |
| 3 | - | - | - |  |  |  |  |  |
| 4 | + | n/a | + | Yes |  |  |  |  |
| 5 | + | n/a | + | Yes |  |  |  | Covid-19 |
| 6 | - | + | + |  | Yes |  |  |  |
| 7 | + | n/a | + |  |  |  |  | (other cause) |
| 8 | + | n/a | + | Yes |  |  |  | Covid-19 |
| 9 | - | + | + |  | Yes |  |  |  |
| 10 | - | - | - |  |  | Yes |  |  |
| 11 | - | - | + | Yes |  |  |  |  |
| 12 | - | - | + |  | Yes |  |  |  |
| 13 | - | - | - |  |  |  |  |  |
| 14 | - | - | - |  |  |  |  |  |
| 15 | - | - | - |  |  |  |  |  |
| 16 | - | - | - | Yes |  |  | Yes |  |
| 17 | - | + | - | Yes |  |  | Yes |  |
| 18 | - | - | + |  |  | Yes |  |  |
| 19 | - | + | - |  |  |  |  |  |
| 20 | + | n/a | + | Yes |  |  | Yes |  |
| 21 | - | - | - |  |  | Yes |  | (other cause) |

The specificity of the RT-LAMP test was 73%. Three cases were identified as positive using RT-LAMP which were negative on RT-PCR, giving a total of 13 patients testing positive on either RT-PCR or RT-LAMP. (Table 3). Of these 3 patients that were positive using RT-LAMP but negative on RT-PCR, patient 11 had a high grade temperature of 38.1°C on D1 of testing, patient 12 had a temperature <36.0°C in the 7 days prior to testing and patient 18 had a temperature of 37.5°C in the 7 days prior to testing (table 2). All three remained well at day 10 with no other explanation for symptoms, such as upper respiratory or urinary tract infections. It is of course possible that the RT-PCR results for one or more of these patients represent false negatives. Of the two patients positive for RT-PCR and negative using RT-LAMP one was contemporaneously symptomatic, and the other was well at the time of testing but had suffered a significant flu-like illness for the 3 weeks prior to Day 0.

**Table 3:** Testing results, comparing RT-LAMP method on deep nasal swabs to Multiplex tandem RT-PCR performed on both pharyngeal and deep nasal swabs. Positive PCR cases are defined as at least one positive test over two days.

|  |  | PCR (repeat) |  |  |
| --- | --- | --- | --- | --- |
|  |  | Positive | Negative | Total |
| LAMP | Positive | 8 | 3 | 11 |
|  | Negative | 2 | 8 | 10 |
| Total |  | 10 | 11 | 21 |
|  |  |  |  | (95% CI) |
| Sensitivity |  | 80% |  | (44%-98%) |
| Specificity |  | 73% |  | (39%-94%) |
| Positive Predictive Value |  | 73% |  | (39%-94%) |
| Negative Predictive Value |  | 80% |  | (44%-98%) |

Many patients in the home had altered vital signs in the week leading up to testing, with 6/11 negative cases, and 8/10 positive cases showing signs, e.g. fevers or reduced oxygen saturations. Low temperatures (<36°C were detected in a minority of COVID PCR positive patients in the absence of fevers (Table 2). The development of cases in the home and testing results are illustrated in Figure 1.

**Figure 1:**
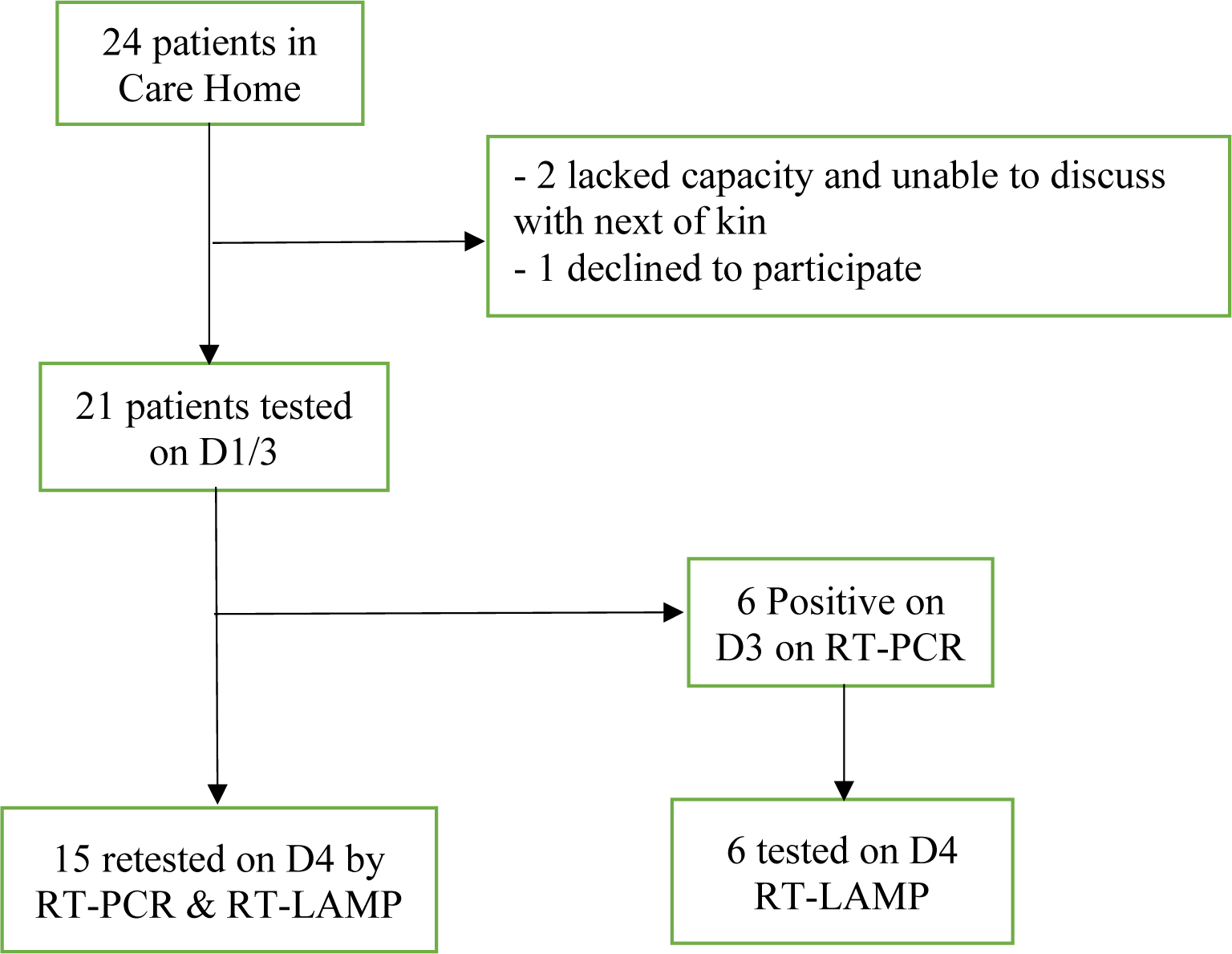
Flowchart of patients.

**Figure 2:**
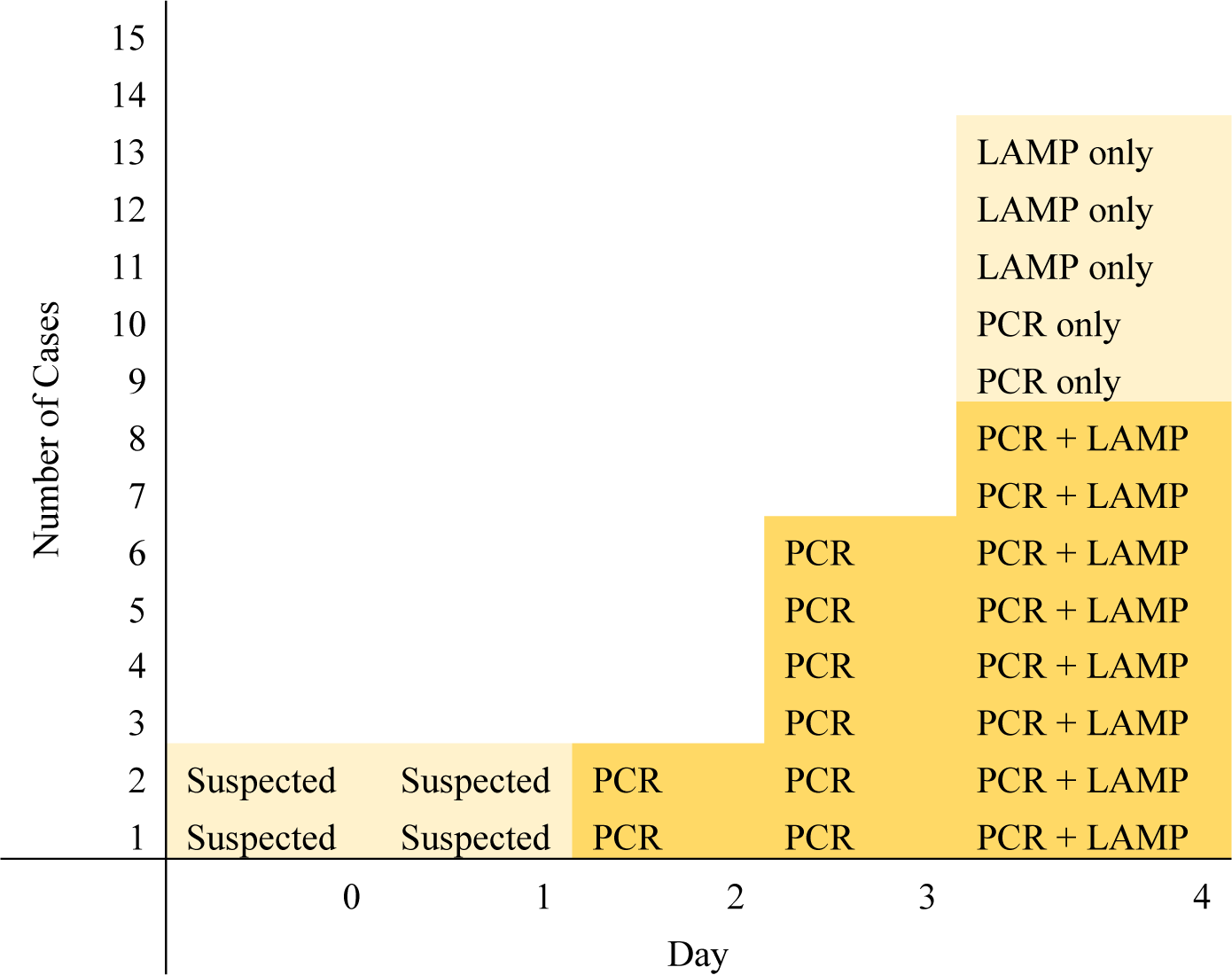
Development of the cases and testing over time.

## Discussion

In a time of global crisis, it is critical data are quickly shared on new testing methods, so that they can be upscaled more rapidly. To this end we present data from 21 patients in a care home tested within days of an outbreak in the home. In this patient group, a single RT-LAMP test had a sensitivity of 80% and a specificity of 73% compared to a “better than gold standard” of two consecutive RT-PCR swabs. This level of sensitivity is clinically workable in a time of crisis, particularly if repeated testing is utilised, and safeguards are put in place to guard against over confidence in negative individuals. It is comparable to other estimates of a single-swab RT-PCR test in our clinical experience and in published pre-prints[7,8]. In addition, it appears nasal swabs are second best to lower respiratory tract derived samples[8]. Combined with the rapid result time, RT-LAMP may have additional clinical utility to standard RT-PCR. The RT-PCR negative, RT-LAMP positive samples may indicate a lack of specificity of the RT-LAMP assay but given that some infected patients are assumed to be have been clinically asymptomatic and given that the RT-LAMP assay used here tests more of the swab eluate than the PCR, these may be real positive results that have missed by the RT-PCR. Further testing and further studies will resolve this issue.

In addition, we found fever >37.3°C, as expected was a common symptom, but hypothermia (T<36.0°C) and desaturation were also noted. The finding of hypothermia is important. It is a recognised symptom of sepsis and the systemic inflammatory response syndrome, particularly in older people^[9]^. However, current PHE and WHO COVID-19 guidelines do not include hypothermia as a symptom. Larger scale studies on prevalence of hypothermia, as well as other non-classical symptoms, would shed more light on the presentation of COVID-19 in institutionalised patients.

Loop-mediated isothermal amplification (LAMP) was developed as a rapid and reliable, cheaper method to amplify from a small amount target sequence at a single reaction temperature, obviating the need for sophisticated thermal cycling equipment^[10]^. Within the past 4-6 weeks, five independent groups have published preprints of submitted manuscripts evaluating novel RT-LAMP testing methods against RT-PCR as gold standard (Table 4). Two of these used only proven PCR-positive throat and nasal swabs and demonstrated sensitivity >97% for RT-LAMP methods targeting the *ORF1ab* gene when compared with gold standard RT-PCR^[11,12]^. Only the study by Yang et al. included swabs from both SARS-CoV-2 positive and negative patients and was thus able to produce both a sensitivity and a specificity. The remaining two groups, both based in in the United States, lacked access to, or clearance to work with, SARS-CoV-2 samples and used either inactivated HIV with synthesised LAMP sequences^[13]^ or other synthesised RT-LAMP sequences ^[14]^. The majority of studies focused on the highly-conserved ORF1ab gene primer, also targeted by the RT-LAMP method used by the MicrosensDx RapiPrep® SARS-CoV-2 method. Our study is the first “real world” study comparing the effectiveness of RT-PCR and RT-LAMP testing in a group of patients at high risk for COVID-19 and represents an important progression to clinical use for this novel SARS-CoV-2 testing method. Of particular note, our standard for comparison was not a single RT-PCR, but two separate swabs for RT-PCR sent on consecutive days, thus representing what could be considered a “better-than gold standard” for comparison.

**Table 4.** Currently available pre-print articles comparing LAMP methods with RT-PCR for COVID-19 detection

| Preprint | Country | Methods | Samples used for validation | Sensitivity of LAMP (for ORF1ab gene) compared with RT-PCR | Specificity of LAMP (for ORF1ab gene) compared with RT-PCR |
| --- | --- | --- | --- | --- | --- |
| El-Tholeth et al. [13] | USA | Two stage isothermal amplification (COVID-19 Penn-RAMP) targetting ORF1ab | No SARS-CoV-2 samples available in USA at time of study so samples with inactivated HIV virus with synthesised LAMP sequences tested. Four positive samples used. | 75% | 100% |
| Lamb et al. [14] | USA | LAMP using unspecified primers | No SARS-CoV-2 samples; synthesised LAMP sequences tested. | Study was not powered to determine sensitivity in a clinical population | N/a |
| Zhang et al. [12] | China | LAMP using ORF1ab, and N gene primers | 6 positive swabs by RT-PCR | 100% | N/a |
| Yu et al. [11] | China | LAMP using ORF1ab gene primers | 43 positive swabs by RT-PCR | 97.6% | N/a |
| Yang et al. [15] | China | LAMP using ORF1ab, E and N gene primers | 208 swabs (17 positive & 191 negative by RT-PCR) | 87.5% (confidence intervals not available) | 99% (confidence intervals not available) |

### Strengths + Limitations

We have been able to perform these tests quickly in a group at high risk for severe disease, and a setting where early identification of infected patients is key to preventing further spread. Many other studies so far have used laboratory samples to estimate efficacy but have been unable to estimate the clinical utility. Swabs were taken by the same clinician, minimising the risk of technical error or observer biases. All RT-LAMP samples were tested in the MicrosensDx laboratory, and RT-PCR in the hospital laboratory, and there was no viral transport medium on the RT-LAMP swabs. Actual Point of Care testing, and or viral medium could be used to optimise performance further but use of dry swabs could ease issues with supply of viral transport media. We are limited by a small sample size, so our estimates have wide confidence intervals. However, they appear to be concordant with other (pre-print) studies on RT-LAMP performed purely on laboratory samples. A further possible limitation of our study is that only upper respiratory samples were used. Additional work is needed to test this method with sputum or bronchiolar lavage specimens, which may improve sensitivity for hospitalised patients. We are also aware that sampling sites were different on Day 3 and 4, however discordant patients had deep nasal samples on Day 4, and this does not appear to have affected our results.

### Generalisability, Implications & Future Research

Use of this rapid test, for example in care home settings could facilitate early identification of cases and enactment of infection control measures as required. We hypothesise that this could significantly reduce spread and subsequent mortality in care home residents, a speculation which could easily be tested if the method was more widely available. The test may also be suitable for use in other community settings such as pharmacies and care agencies, as well as emergency departments, and prisons or residential settings for homeless people where rapid diagnosis is also important. An area of global concern is COVID-19 spread in developing countries, where reported cases are currently low but are likely to increase over the coming weeks and months. Inexpensive PoC testing that is not dependent on skilled and centralised technicians will be vital for the less well-resourced countries and economies. However, further evaluation in these settings would be advised to replicate its effectiveness.

## Conclusion

There is urgent need of a rapid, robust and cost-efficient point-of-care test that can be used in care homes, community settings and away from centralised large-scale laboratories, without the need for skilled technicians. Magnetic bead capture and RT-LAMP amplification and testing for SARS-CoV-2 was found to be promising, rapid, easy to use and to work equivalently to standard multiplex tandem PCR methods. Definitive studies to evaluate this method in larger cohorts are underway. RT-LAMP has the potential to transform COVID-19 detection, bringing rapid and accurate testing to the PoC.

## Data Availability

Data used in this paper will be published in supplementary files in the peer-reviewed publication.

## Other Information

CJS is supported by HEFCE funding. CS and SW are employees of MicrosensDX Ltd. Testing was provided free of charge by MicrosensDx. PCR tests were performed as part of routine clinical care. No other funding was sought for this study. Other authors report no conflict of interest.

## Ethical Considerations

This project was a clinical service improvement and as such did not require REC approval. All capacitous participants and relatives in case of non-capacitous were appraised of the project and given the opportunity to not take part. This decision had institutional oversight from the offices of the Integrated Care Medical Director and Research and Development, Guy’s & St Thomas’ NHS Trust, and the Clinical Academic Group for Medicine and Therapies, King’s Health Partners.

